# Longitudinal associations between childhood socioeconomic position and adolescent eating disorder symptoms: findings from the ALSPAC cohort

**DOI:** 10.1101/2025.01.12.25320417

**Authors:** Jane S. Hahn, Eirini Flouri, Amy Harrison, Glyn Lewis, Francesca Solmi

**Affiliations:** UCL Division of Psychiatry; UCL Institute of Education

## Abstract

**Background:** The recorded incidence of eating disorders is elevated in people from higher socioeconomic positions, but self-reported eating disorder symptoms are more common in those from lower socioeconomic positions in cross-sectional studies. Longitudinal studies investigating the role of different socioeconomic indicators in the aetiology of a range of eating disorder symptoms might avoid biases associated with the use of clinical samples but have multiple limitations. We aimed to investigate the association between early childhood socioeconomic indicators and eating disorder symptoms across adolescence.

**Method:** We used data from the Avon Longitudinal Study of Parents and Children. Parental income, education, occupation, and financial hardship were reported by mothers between 32 weeks of gestation and 47 months postpartum. Area-level deprivation was derived from the Office for National Statistics indicators linked to the participant’s residential post code at 32 weeks gestation. Outcomes were disordered eating (restrictive eating, binge eating, and purging), weight and shape concerns, and body dissatisfaction at age 14, 16, and 18 years. To model these associations, we used univariable and multivariable multilevel logistic and linear regression, and linear regression models.

**Findings:** The sample included 7,824 participants (48.9% females; 3.8% minoritised ethnic background). Greater financial hardship was associated with increased odds of disordered eating (Odds Ratio= 1·06, 95%CI 1·04 to 1·10) and greater weight and shape concerns (Mean difference[MD] = 0·02, 95%CI 0·01 to 0·04) and body dissatisfaction (MD= 0·22 95%CI 0·06 to 0·37). Lower parental education was associated with 1.80 higher odds of disordered eating (95% CI 1·46 to 2·23).

**Interpretation:** Our findings point to potential socioeconomic inequalities in eating disorder identification in clinical settings, which need to be understood and addressed. Reducing population-level socioeconomic inequalities could also aid eating disorder prevention.

**Funding source:** Mental Health Research UK, Wellcome Trust, UK Medical Research Council

## Background

Globally, socioeconomic deprivation is a major determinant of poor mental and physical health in children. ^1, 2^ In the UK, one in three children live in poverty, with increasing proportions living in extreme poverty. This raises concerns about the long-term wellbeing of a large portion of young people, ^3^ as children from the most deprived households have three-fold higher prevalence of mental health problems such as depression and anxiety compared to those living in the least deprived households. ^1, 2^

In contrast, it is often believed that eating disorders are more common in people with families from higher socioeconomic positions, ^2, 4, 5^ but evidence supporting this association is mixed. Most longitudinal register-based studies, where diagnoses are derived from clinical records, find a higher incidence of eating disorders in people whose parents had higher income and education, and who lived in more affluent areas . ^2, 4^ Conversely, cross-sectional^6^ and longitudinal population^7-14^ studies either find no differences in the distribution of self-reported eating disorder symptoms by parental socioeconomic position^8, 14^ or show increased risk of these symptoms in people from more deprived backgrounds. ^6-13^

Nevertheless, the literature has several limitations which limit our understanding of the association between childhood socioeconomic position and adolescent eating disorders. Findings from register-based studies may be affected by selection bias if people from more deprived backgrounds experience barriers in accessing eating disorder services. ^15 6^ Longitudinal studies either used a long follow-up spanning into adulthood^2, 4, 12, 13^ or, when focusing on young people, did not include the peak time of eating disorder symptom onset, ^7-9, 11, 14^ which could affect findings if early onset cases are underpinned by different aetiological mechanisms. ^16^ Most studies adjusted their analyses for factors which are potentially on the causal pathway between family socioeconomic position and offspring eating disorder, such as adverse life experiences^13^ or offspring’s BMI, ^8-10, 12, 13^ which can bias results. Finally, existing studies used either a single measure of socioeconomic position (often education) or composite indices. As policies needed to address specific socioeconomic domains might differ, exploring a wide range of socioeconomic indicators can help inform future preventative strategies. Therefore, we investigated here the longitudinal association between parental income, occupation, education, financial hardship, and area-level deprivation in early childhood and adolescent eating disorder symptoms in a large UK general-population cohort.

## Methods

### Sample

We used data from the Avon Longitudinal Study of Parents and Children (ALSPAC), a birth cohort study which recruited 14,541 pregnant women in the former region of Avon (UK) with expected delivery dates from 1^st^ April 1991 to 31^st^ December 1992. Of these pregnancies, 14,062 (96·1%) resulted in live births and 13,988 (93·8%) children were alive at one year. ^17, 18^ In this study, we included children from this original sample who had data available on all the exposures. In the case of twins, we retained one child at random to avoid potential over-estimation of associations due to clustering of environmental and genetic risk. The research ethics committee at the University of Bristol and the ALSPAC Ethics and Law Committee provided ethical approval for the study.

### Outcomes

We used three different outcomes capturing behavioural (disordered eating) and cognitive (weight and shape concerns, body dissatisfaction) symptoms of eating disorders.

Disordered eating was measured at 14, 16, and 18 years through self-report of binge eating, purging (i.e. self-induced vomiting or laxative use), excessive dieting, and fasting in the previous 12 months using a set of modified questions from the Youth Risk Behaviour Surveillance System questionnaire. ^19^ In line with previous studies, we defined disordered eating as a binary variable indicating whether adolescents reported any of these behaviours. ^20^ We also used these behaviours individually as a secondary outcome to investigate their specific associations with socioeconomic position. See supplement (p.1).

Body dissatisfaction was self-reported at age 14 years, using the body satisfaction scale. ^21^ Adolescents reported satisfaction on 11 aspects of their body (e.g., weight and figure) on a four-point Likert scale ranging from extremely satisfied (5) to extremely dissatisfied (1). Scores were reverse coded and summed. The total score ranged from 11 to 55; higher scores indicated greater body dissatisfaction.

Weight and shape concerns were self-reported by adolescents at 14 and 18 years using two items from the McKnight Risk Factor survey^22^ asking whether adolescents were “happy with the way their body looks in the past year” and whether “weight made a difference to how they felt about themselves in the past year.” Responses were scored on a four-point Likert scale ranging from 0 (‘very unhappy’/’a lot’) to 3 (‘very happy’/’not at all’). We reverse coded and summed these scores. Scores ranged from 0 to 6; higher scores indicated greater concerns.

### Exposures

We derived highest parental occupation (professional, managerial, skilled non-manual, skilled manual, and semi-skilled/unskilled manual) and highest parental education (university-degree, A-level, compulsory education) from individual paternal and maternal measures as reported by the mother at 32 weeks of gestation. If either parent had missing data on these variables, or in cases of single-parent households, we used data on the available parent.

At this time point, mothers were also asked how difficult they were finding it to afford food, heating, clothing, rent or mortgage, and items for their baby. Answers were scored on a four-point Likert scale ranging from “not difficult” (0) to “very difficult” (3). From these we derived a continuous score ranging from 0 to 15 where higher scores represented greater financial hardship.

Mothers reported weekly family income when the study child was 33 months and 47 months. Parental income was averaged across these time points and weighted by number of people within the household according to their age and estimated housing benefits using the Organisation for Economic Co-operation and Development modified scale and split into fifths. ^23^

Mothers also provided residential post-codes throughout gestation. These were mapped onto enumeration district codes (small census areas) and linked to 1991 Census data Townsend deprivation index scores^24^ which we subsequently standardised. Higher values indicate greater deprivation (supplement p.3).

### Confounders

We identified confounders based on literature-informed a-priori assumptions and using direct acyclic graphs to model our assumptions.

In main analyses, we only mutually adjusted each exposure for all other indicators of socio-economic position given their interconnectedness, which is often intergenerational. We did not include child characteristics (e.g. sex) as those are unrelated to family socioeconomic status or on the causal pathway between the latter and eating disorder risk (e.g. mental health difficulties). We also hypothesised that maternal characteristics (e.g. marital status, age at child’s birth, and history of eating disorder and depression) could be on the causal pathway between exposures and outcomes, as socioeconomic position in pregnancy could reflect earlier socioeconomic position and this could affect subsequent maternal socioeconomic indicators. .

In sensitivity analyses, however, we tested competing causal assumptions and confounding structures. First, we hypothesised that some socioeconomic indicators could affect others, for instance that education could affect subsequent income (**Supplemental Figure S2**, p.5). Second, we hypothesised that maternal characteristics^25, 26^ could affect subsequent socioeconomic position; for instance, maternal history of eating disorders could affect the mother’s educational attainment since peak age onset of eating disorders coincides with adolescence (**Supplemental Figure S3**, p.6). In sensitivity analyses, given the small number of ethnic minority participants, we also additionally adjusted analyses for child’s ethnicity as a proxy of parental ethnicity, as ethnic minorities face additional barriers which can affect their socioeconomic status and eating disorder risk (**Supplement**, p.8). ^27^

### Data analysis

The study protocol was pre-registered on Open Science Framework (https://osf.io/hsg85/). We reported any small protocol deviations in the supplemental material (p.9).

We described sample characteristics overall and by levels of exposures using frequencies with proportions and means with standard deviations. For participants with complete exposure data, we compared the distribution of exposures and confounders between participants who had missing outcome measures.

To investigate the association between each socioeconomic indicator and eating disorder symptoms, we used univariable and multivariable multilevel logistic (any and each individual disordered eating) and linear (for weight and shape concerns) regression models with time of outcome assessment nested within individuals. First, we ran an unconditional model only including a mean-centred indicator of age at outcome measurements to describe how the latter changed across adolescence. For disordered eating, where we had three measurements available, we also added a quadratic term for age to test for non-linear associations with age, retaining this in the models if there was evidence of an association.

Subsequently, we ran a univariable model for each exposure and a multivariable model, adjusting each exposure for all other indicators of socioeconomic position. In the fully adjusted model, we subsequently included an interaction between each exposure and age to investigate whether there were differential associations with the exposure based on timing of outcome measurement. We stratified results by age where we found evidence of an interaction. To investigate associations with body dissatisfaction at age 14 years, we used univariable and multivariable linear regression models mutually adjusting each socioeconomic indicators for all other indicators. We imputed missing confounder and outcome data using multiple imputation by chained equations for participants with complete data on exposure. We imputed 50 datasets on the assumption that the data were missing at random using all the variables included in the final models and auxiliary variables (**Supplement**, p.10).

We ran four sets of sensitivity analyses. First, we further adjusted the main multivariable models for maternal marital status, and history of eating disorders and depression. Second, we re-ran the main multivariable models adjusting parental occupation for parental education; family income for parental occupation and education; and financial hardship for family income and highest parental occupation and education. We also adjusted all the main analyses for participants’ ethnicity. The rationale for all these analyses is explained in the ‘*confounders’* section. To explore how ethnicity may confound our associations, we adjusted all the main analyses for ethnicity to see how it may affect the effect sizes and estimates from the main analyses. To explore whether missing data patterns affected our effect sizes and estimates, we ran all our main analyses in a sample of participants with complete exposures and outcome (for body dissatisfaction) or at least one time-point of outcome measurement available (for disordered eating and weight and shape concerns).

## Results

### Sample characteristics

From the total sample of ALSPAC children alive at one year (n=13,988), 7,824 (55.9%) children had complete data on all exposures after removing one twin and were thus included in the analytical sample.

A large proportion of participants’ parents had a managerial occupation (43.5%) and compulsory education as their highest educational qualification (40.0%), and were in the highest fifth of income categories (21.8%). Most families did not experience financial hardship (76.4%) and lived in areas of low deprivation (73.7%) during pregnancy. (**Table 1**)

**Table 1:**
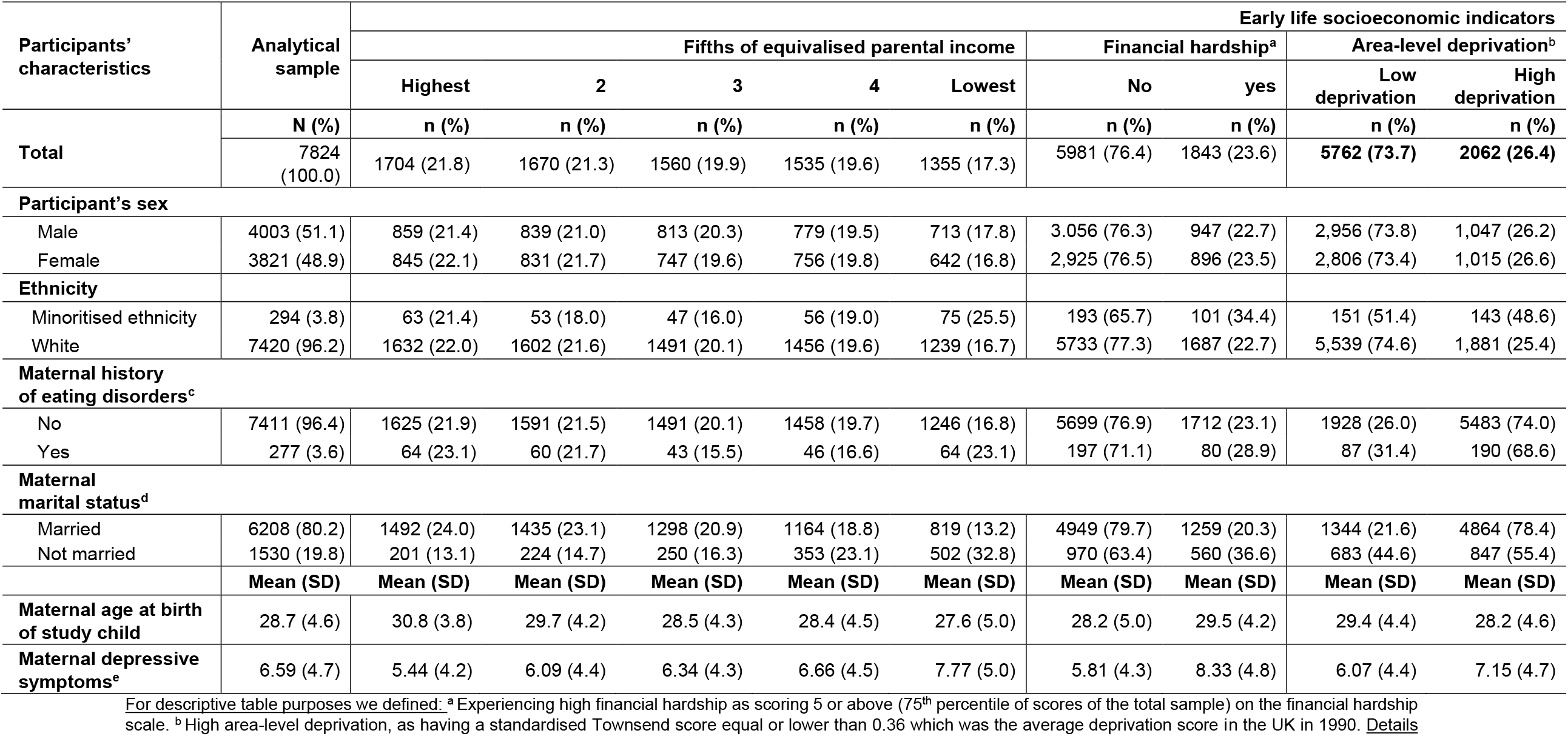
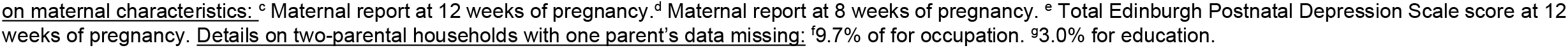
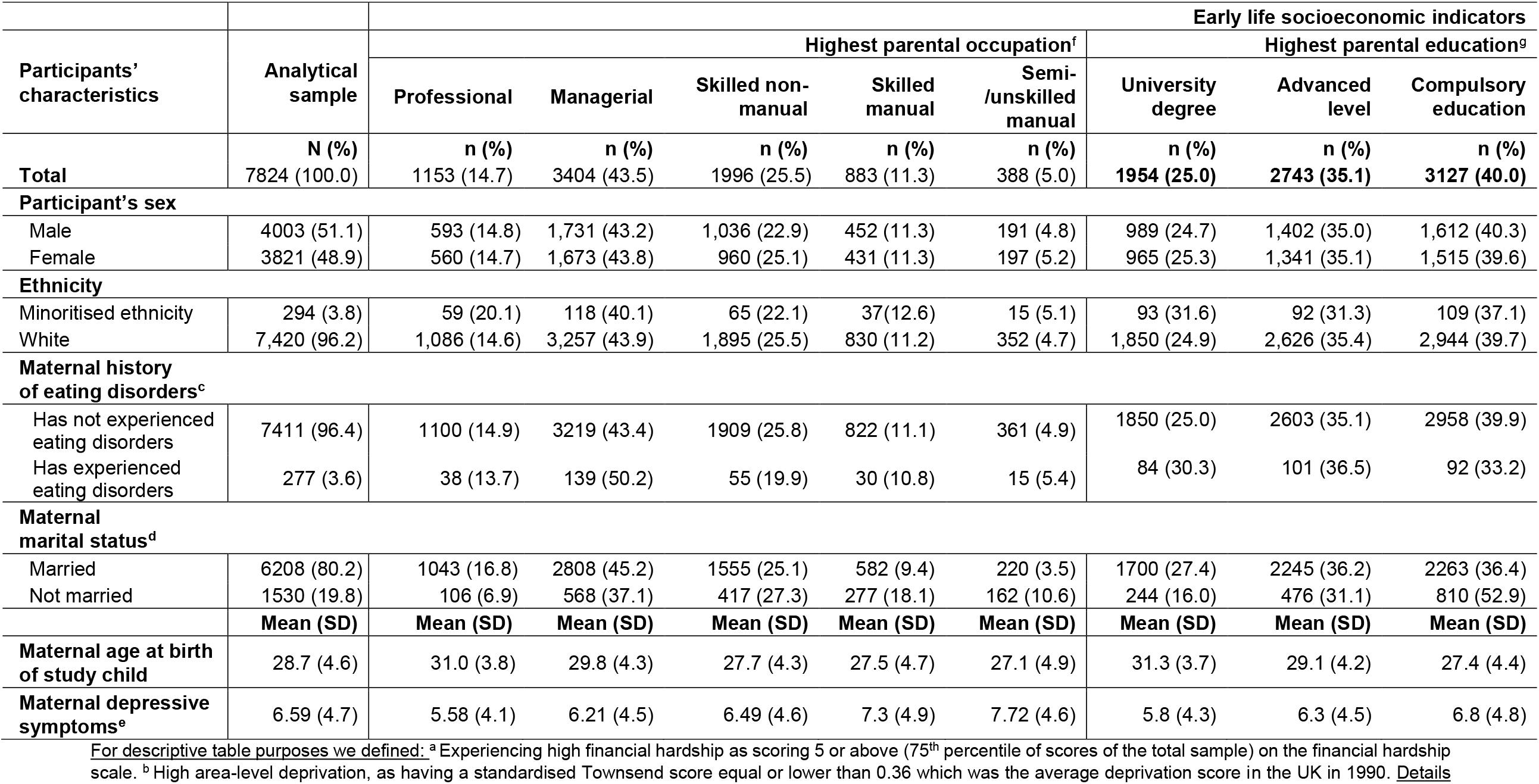
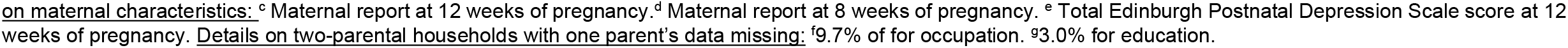
Sample characteristics

**Table 3:** Univariable and multivariable Multilevel logistic and linear regression models for any behavioural eating disorder symptoms and weight and shape concerns at age 14, 16, and 18 according to parental socioeconomic position. Sample based on participants with complete exposure data (N=7824)

| Parental socioeconomic position | Disordered eating |  | Weight and shape concerns |  |
| --- | --- | --- | --- | --- |
|  | Univariable model<br>Odds ratios<br>(95% CI), p-value | Multivariable model<br>Odds ratios<br>(95% CI), p-value | Univariable model<br>Mean difference<br>(95% CI), p | Multivariable model<br>Mean difference<br>(95% CI), p |
| **Highest parental education** |  |  |  |  |
| University degree | Ref | Ref | Ref | Ref |
| A -level | 1.35 (1.09 to 1.68), 0.007 | 1.31 (1.03 to 1.66), 0.030 | 0.07 (-0.02 to 0.15), 0.124 | 0.02 (-0.07 to 0.12), 0.673 |
| Compulsory education | 1.80 (1.46 to 2.23), <.0001 | 1.64 (1.24 to 2.16), 0.001 | 0.08 (-0.002 to 0.18), 0.055 | 0.02 (-0.09 to 0.14), 0.701 |
| **Highest parental occupation** |  |  |  |  |
| Professional | Ref | Ref | Ref | Ref |
| Managerial | 1.27 (1.00 to 1.62), 0.046 | 1.05 (0.81 to 1.36), 0.728 | 0.10 (0.01 to 0.28), 0.028 | 0.08 (-0.03 to 0.19), 0.144 |
| Skilled non-manual | 1.54 (1.17 to 2.02), 0.002 | 1.09 (0.78 to 1.51), 0.610 | 0.13 (0.03 to 0.22), 0.012 | 0.08 (-0.04 to 0.20), 0.197 |
| Skilled manual | 1.54 (1.08 to 2.19), 0.015 | 0.99 (0.65 to 1.51), 0.966 | 0.12 (-0.04 to 0.27), 0.141 | 0.05 (-0.13 to 0.23), 0.611 |
| Semi-skilled/unskilled | 2.09 (1.32 to 3.34), 0.002 | 1.26 (0.75 to 2.11), 0.378 | 0.18 (-0.05 to 0.40), 0.123 | 0.09 (-0.15 to 0.35), 0.434 |
| **Fifths of equivalised family income** |  |  |  |  |
| Highest 20% | Ref | Ref | Ref | Ref |
| 2 | 0.99 (0.77 to 1.30), 0.990 | 0.84 (0.65 to 1.10), 0.211 | 0.02 (-0.07 to 0.12), 0.628 | -0.01 (-0.11 to 0.09), 0.792 |
| 3 | 1.08 (0.82 to 1.42), 0.595 | 0.79 (0.59 to 1.04), 0.096 | 0.07 (-0.04 to 0.19), 0.214 | 0.006 (-0.12 to 0.13), 0.925 |
| 4 | 1.35 (1.04 to 1.74), 0.024 | 0.88 (0.66 to 1.16), 0.355 | 0.10 (-0.01 to 0.21), 0.085 | -0.0002 (-0.12 to 0.12), 0.996 |
| Lowest 20% | 1.34 (1.01 to 1.79), 0.042 | 0.75 (0.55 to 1.03), 0.077 | 0.06 (-0.07 to 0.18), 0.368 | -0.08 (-0.23 to 0.06), 0.268 |
|  | OR (95% CI), p | OR (95% CI), p | Coefficient (95% CI), p | Coefficient (95% CI), p |
| **Financial hardship score** | 1.07 (1.04 to 1.10), <.0001 | 1.06 (1.03 to 1.09), <.0001 | 0.02 (0.01 to 0.04), 0.001 | 0.02 (0.01 to 0.04), 0.003 |
| **Standardised area-level deprivation score** | 1.05 (1.02 to 1.09), 0.002 | 1.03 (0.99 to 1.07), 0.067 | 0.01 (0.001 to 0.03), 0.035 | 0.01 (-0.003 to 0.02), 0.122 |

**Table 4:** Linear regression model for body dissatisfaction at age 14 according to parental socioeconomic position. Sample based on participants with complete parental socioeconomic data and imputed eating disorder outcomes (N=7824)

| Parental socioeconomic position indicators | Body dissatisfaction |  |
| --- | --- | --- |
|  | Univariable model<br>Mean difference (95% CI), p-value | Multivariable model<br>Mean difference (95% CI), p-value |
| **Highest parental education** |  |  |
| University degree | Ref | Ref |
| A -level | 0.23 (-0.94 to 1.41), 0.694 | -0.03 (-1.12 to 1.06), 0.963 |
| Compulsory education | 1.08 (-0.08 to 2.23), 0.069 | 0.57 (-0.65 to 1.79), 0.355 |
| **Highest parental occupation** |  |  |
| Professional | Ref | Ref |
| Managerial | 0.34 (-1.02 to 1.70), 0.620 | 0.01 (-1.40 to 1.43), 0.984 |
| Skilled non-manual | 0.77 (-0.72 to 2.25), 0.307 | 0.11 (-1.50 to 1.73), 0.889 |
| Skilled manual | 1.49 (-0.69 to 3.66), 0.177 | 0.48 (-1.89 to 2.85), 0.686 |
| Semi-skilled/unskilled | 0.93 (-1.95 to 3.81), 0.523 | -0.27 (-3.25 to 2.72), 0.857 |
| **Fifths of equivalised family income** |  |  |
| Highest 20% | Ref | Ref |
| 2 | 0.61 (-0.78 to 2.01), 0.384 | 0.31 (-1.04 to 1.67), 0.644 |
| 3 | -0.35 (-2.09 to 1.39), 0.690 | -0.94 (-2.69 to 0.81), 0.285 |
| 4 | 2.17 (0.45 to 3.91), 0.014 | 1.27 (-0.48 to 3.01), 0.151 |
| Lowest 20% | 0.72 (-1.07 to 2.50), 0.427 | -0.61 (-2.58 to 1.37), 0.540 |
|  | Coefficient (95% CI), p value | Coefficient (95% CI), p value |
| **Financial hardship score** | 0.24 (0.10 to 0.39), 0.001 | 0.22 (0.06 to 0.37), 0.007 |
| **Standardised Area-level deprivation score** | 0.13 (-0.03 to 0.29), 0.116 | 0.07 (-0.09 to 0.22), 0.395 |

The distribution of participants in terms of sex assigned at birth was comparable across all socioeconomic position indicators. Parents of children from minoritised ethnic background had lower income, experienced more financial hardship, and lived in higher deprivation areas. A greater proportion of participants with unmarried mothers reported more deprivation across all indicators. Average levels of maternal depressive symptoms were progressively higher and mean maternal age lower in categories denoting more deprived backgrounds. (**Table 1**)

### Missing data

Outcome measurements were more commonly missing among participants from more deprived backgrounds across all socioeconomic indicators as well as in those with younger and single mothers as well as mothers with greater depressive symptoms. (**Supplemental Table S1**, p.11-12)

### Early-life socioeconomic position and adolescent disordered eating

In unconditional models, there was evidence of a linear (odds ratio [OR]=1.37, 95% confidence intervals [CI] 1.30 to 1.44, p<.001) and non-linear (OR=0.92, 95% CI 0.89 to 0.96, p<.001) association between age and disordered eating and therefore both age indicators were retained in subsequent analyses. (**Supplemental Tables S2, S3**, p.13)

In univariable models, participants whose parents had only completed compulsory education (OR=1·80, 95% CI 1·46 to 2·23), had a semi-skilled/unskilled occupation (OR = 2·09, 95% CI 1·32 to 3·34), and whose income was in the lowest fourth (OR = 1·35, 95% CI 1·04 to 1·74) and fifth of income distribution (OR = 1·34, 95% CI 1·01 to 2·79) had higher odds of experiencing disordered eating compared to adolescents whose parents had university-level education or a professional occupation, and who were in the highest fifth of income distribution respectively. A one-point increase in financial hardship score (OR=1·07, 95% CI 1·04 to 1·10) and a standard deviation increase in area-level deprivation (OR=1·05, 95% CI 1·02 to 1·09) were associated with higher odds of experiencing disordered eating in adolescence.

After adjusting for the remaining socioeconomic indicators, there was still strong evidence that adolescents whose parents only had compulsory education had higher odds of disordered eating compared to adolescents whose parents had university-level education (OR = 1·64, 95% CI 1·24 to 2·16) and that a one-point increase in financial hardship score was associated with 1.06 higher odds (95% CI 1·03 to 1·09) of disordered eating. Higher area-level deprivation was also still associated, albeit weakly, with increased odds of disordered eating in the multivariable model (OR=1.03, 95% CI 0.99 to 1.07) but there was no evidence of an association for lower parental occupation and income.

When investigating interactions between each of the exposure and age of outcome measurements, we found some evidence of an interaction between income and age (p-value for interaction=0.032). Participants from lower income brackets had higher odds of experiencing disordered eating when they were younger (age-stratified results in supplemental Table S4 p.14).

Analyses of individual disordered eating revealed similar patterns as the main analysis, but we found that lower education and greater financial hardship were associated with increased odds of restrictive eating and higher area-level deprivation with increased odds of binge eating and purging (Supplemental Table S5, p.15-16).

### Early-life socioeconomic position and adolescent weight and shape concerns and body dissatisfaction

In the unconditional models, there was evidence of a linear association (coefficient=0.09, 95% CI 0.07 to 0.11, <.0001) between age and weight and shape concerns, so we included this variable in subsequent analyses. (**Supplemental Tables S2, S3**, p.13)

In univariable models, there was no evidence of an association between lower parental income, occupation status, and educational attainment and both weight and shape concerns, and body dissatisfaction (Table 2). A one-point increase in parental financial hardship score was associated with a 0·02-point increase (95% CI 0·01 to 0·04) in weight and shape concern score and a 0·24-point increase (95% CI 0·10 to 0·39) in body dissatisfaction score. These associations remained unchanged once adjusting for all other socioeconomic indicators (coefficient of increase for one-point increase in financial hardship score in weight shape concerns score: 0·02; 95% CI 0·01 to 0·04 and body dissatisfaction score: 0·22; 95% CI 0·06 to 0·37).

In univariable models, a one-standard deviation increase in area-level deprivation was associated with a 0·01-point increase (95% CI 0·001 to 0·03) in weight and shape concern scores, but evidence of this association was no longer present once adjusting for all other indicators. We did not observe evidence of an interaction between socioeconomic position and age of outcome measurement for weight and shape concerns.

### Sensitivity analyses

Sensitivity analyses (i) adjusting for *ad hoc* socioeconomic indicators (Supplemental Table S5, S6, p.17-19), (ii) including maternal characteristics (Supplemental Table S7, S8, p.22-23) and (iii) child ethnicity (Supplemental Table S11, S12, p.24-26), or (iv) restricted to participants with complete data (Supplemental Table S9, S10, p.22-23) yielded comparable estimates to those observed in the main analyses, albeit, in some cases, wider confidence intervals.

## Discussion

In this study, we found that participants from more deprived backgrounds experienced greater eating disorder symptoms throughout adolescence. More severe financial hardship was associated with increased risk of disordered eating, weight and shape concerns, and body dissatisfaction. Lower parental educational attainment was strongly associated with increased odds of offspring’s disordered eating in adolescence.

### Limitations

Despite its large sample, longitudinal design, and nuanced measurement of both outcome and exposures, our study has some limitations. In ALSPAC, there are high levels of attrition among respondents from lower socioeconomic backgrounds. This might have biased our results if those participants have differential risk of eating disorder symptoms. We addressed this limitation by imputing missing outcome data for those with complete exposures and were reassured to observe comparable results across complete record and imputed analyses.

Using the data available for parental occupation and educational attainment may have introduced bias in our associations if missingness patterns relate to both the exposure and the outcome. 9.7% of two-people households had data missing from one parent for occupation and 3.0% for education.

Our measurement of restrictive eating behaviours may not accurately capture extreme restrictive behaviours, typical symptoms of anorexia nervosa, as these are uncommon in general-population samples. Our outcome might have instead captured young people with more common restrictive eating behaviours which could have different patterns of associations with socioeconomic status.

Previous research has shown that genetic susceptibility for a number of psychiatric conditions, including anorexia nervosa, is associated with increased probability of being born in more deprived environments – possibly as a result of intergenerational drift. ^24^ Although we adjusted our models for maternal mental health difficulties in sensitivity analyses, we were unable to robustly account for the potential for genetic confounding as polygenic risk scores currently explain limited phenotypical variance^28^ and the sample did not allow other genetically-informed designs.

### Interpretation of findings and comparison with previous literature

Previous general-population longitudinal investigations have reported an association between socioeconomic position and self-reported eating disorder symptoms in early adolescence. ^7-13^ We expand on those findings by showing that the association between deprivation and eating disorder symptoms extends to the full adolescent period, suggesting persistent effects of early-life deprivation.

Greater financial difficulties and lower parental educational attainment have the strongest associations with the full spectrum of eating disorder symptoms, but mechanisms underpinning these associations remain unexplored. It is possible that putative risk factors for eating disorders more commonly observed in those from lower socioeconomic positions, such as higher child BMI, increased food insecurity, and greater experience of childhood adversities, ^9^ might explain these associations.

The lack of evidence for an association between low parental education and cognitive symptoms of eating disorders is surprising as these usually precede onset of behavioural symptoms. ^29^ It is possible that we might not have been able to observe those associations due to low statistical power, although other studies with smaller sample sizes have shown an association between lower socioeconomic position and cognitive eating disorder symptoms. ^10-12, 14^ However, if these findings were true, they could suggest different risk mechanisms between parental education, disordered eating and cognitive eating disorder symptoms.

On the other hand, our results stand in stark contrast with those of register-based studies. ^2, 4^ Anorexia nervosa is often over-represented in clinical samples. ^4^ Therefore, if anorexia nervosa has a different pattern of association with socioeconomic position, we might not have been able to capture this using our outcome measure. However, register-based studies also find an association between high socioeconomic position and higher incidence of diagnosed bulimia nervosa and eating disorder not otherwise specified^4^ – which we should have captured more accurately with our outcome measures. We therefore hypothesise that the discrepancy between the socioeconomic patterning of clinical diagnoses and self-reported symptoms might be explained by the presence of inequalities in identification of eating disorders and access to services rather than measurement issues.

People from more deprived backgrounds generally experience greater difficulties in accessing healthcare. However, eating disorders are one of the few conditions where an association with deprivation is either not observed or reversed when using clinical registers, suggesting that there might be eating disorder-specific barriers in access to care. First, people with eating disorders from more deprived backgrounds could be less likely to seek help due to the internalization of stigmatising beliefs that eating disorders are a “disease of affluence”. Supporting this hypothesis, a US-based study found that participants from less affluent backgrounds experiencing eating disorder symptoms were less likely to perceive need for treatment. ^30^ Second, those with higher BMI are less likely to receive consultation for eating disorders, ^31^ which might limit referrals for adolescents from lower socioeconomic positions who are on average more likely to have higher BMI. ^32^

### Implications of findings

Identifying and addressing existing barriers that might prevent young people from deprived backgrounds from accessing eating disorder services should be a research and a policy priority. In the UK, medical students receive either minimal or no eating disorder training. ^33^ Provision of comprehensive training might facilitate identification of a broader spectrum of eating disorder presentations in primary care, particularly in populations who are more likely to be missed. Lastly, our findings add to the extensive evidence base calling for a reduction in socioeconomic inequalities as part of population-wide mental health prevention strategies.

## Contributors

**Jane Hahn**: conceptualization, methodology, formal analysis, writing – original draft; **Amy Harrison**: conceptualization, methodology, writing – review & editing, supervision, and funding acquisition; **Eirini Flouri**: conceptualization, methodology, writing – review & editing, supervision; **Glyn Lewis**: conceptualization, methodology, writing – review & editing, and supervision; **Francesca Solmi**: conceptualization, methodology, writing – review & editing, supervision, project administration, and funding acquisition.

## Data Availability

All data produced in the present study are available upon reasonable request to the authors

## Acknowledgements

We are extremely grateful to all the families who took part in this study, the midwives for their help in recruiting them, and the whole ALSPAC team, which includes interviewers, computers and laboratory technicians, clerical workers, research scientists, volunteers, managers, receptionists, and nurses. The UK Medical Research Council and Wellcome (Grant ref: 217065/Z/19/Z) and the University of Bristol provide core support for ALSPAC. This publication is the work of the authors will serve as guarantors for the contents of this paper.

