## Supplement for "Longitudinal associations between childhood socioeconomic position and adolescent eating disorder symptoms: findings from the ALSPAC cohort"

Jane Sungmin Hahn, MSc

UCL Division of Psychiatry, Maple House, 6th floor Wing A,

149 Tottenham Court Rd, W1T 7NF

London, UK.

**Outcome**

To assess the presence of binge eating, adolescents were asked "during the past year, how often did you go on an eating binge?". Eating binge was defined as eating "an amount of food that most people would consider to be very large, in a short period of time." Possible responses were "never", "less than once a month", "1-3 times a month", "once a week", and "more than once a week". As a follow-up question, adolescents were asked whether they felt out of control during these episodes of overeating. Responses included "no", "yes, sometimes", and "yes, usually". Adolescents were classified as having experienced binge eating if they reported overeating at least "1-3 times a month", and if they answered "yes sometimes" or "yes usually" in the follow-up question on loss of control.

To assess the presence of purging, adolescents were asked "during the past year, how often did you make yourself throw up (vomit) to lose weight or avoid gaining weight?". Possible responses were "never", "less than once a month", "1-3 times a month", "once a week", "2-6 times a week", and "everyday". Adolescents were also asked whether they had used laxatives to lose or avoid gaining weight during the past year at 14 years of age and whether they used laxatives or other tablets or medications to lose or avoid gaining weight in the past year at 16 and 18 years of age. The responses were formatted the same as the question on vomiting. Respondents were classified as having experienced purging behaviours if they answered that they self-induced vomit or used laxatives/other tablets or medication to lose weight at least "1-3 times a month".

Adolescents were asked "during the past year, how often did you fast (not eat for at least a day) to lose weight or avoid gaining weight?". We classified adolescents as fasting if they indicated that they fasted at least "1-3 times a month" in the past year from responses "never", "less than once a month", "1-3 times a month", "once a week", and "more than once a week". Adolescents were also asked "during the past year, did you go on a diet to lose weight or keep from gaining weight" with possible responses including "never", "a couple of times", "several times", "often", and "always on a diet". Adolescents were classified as extreme dieting if they answered, "always on a diet" or "often". We coded the presence of restrictive eating behaviours based on respondents who fit these two criteria and who indicate their binge eating frequency as "less than once a month" because those who have restrictive subtypes of eating disorders still engage in occasional binge eating behaviour.[1]

We measured body dissatisfaction using the body dissatisfaction scale which was measured when the adolescents were 14 years old.[2] Adolescents rated their satisfaction with nine body parts including weight, figure, stomach, waist, thighs, buttocks, hips, legs, and hair. Female adolescents were additionally asked about satisfaction with 'breasts' whereas males were asked about satisfaction with 'body build'. Potential responses included "extremely satisfied" (1), "moderately satisfied" (2), "can't decide" (3), "moderately dissatisfied" (4), "extremely dissatisfied" (5). Responses that indicated "can't decide" were coded as missing. Responses that indicated that the body part was "not an issue" were coded the same as those who were "extremely satisfied" with the body part. The total score ranged from 11 to 55 with a higher score indicating a higher level of dissatisfaction.

Weight and shape concerns were measured when the respondents were 14 and 18 years old using two questions from the McKnight Risk Factor survey[3]: 'in the past year' 1) 'how happy have you been with the way your body looks?'; and 2) 'In the past year, how much has your weight made a difference to how you feel about yourself?'. Adolescents could respond on a

Likert scale ranging from 0 (‘very unhappy’/’’a lot’) to 3 (‘very happy’/’not at all’). We added these two items and reversed the scores so higher scores indicated more concern over body weight and shape.

**Exposures**

Family income

Mothers were asked about net family income when the children were 33, 47, and 85 months old. In the questionnaire the net income of the family was recorded in five income bands (<£100, £100 to £199, £200 to £299, £300 to £399, >£400 per week). Income was averaged at 33 months and 47 months, equivalised [4] – i.e., weighed by number of people within the household according to their age and estimated housing benefits – by using the OECD modified scale and split into fifths.[5]

Highest parental occupation

Mothers reported their occupation and that of their partner via postal questionnaire at 32 weeks gestation. From these, we derived a single highest parental occupation variable. We grouped parental occupation from standard categories measured by ALSPAC based on the National Statistics Socioeconomic Classification (unskilled, semi-skilled manual, skilled manual, skilled non-manual, managerial, and professional) into professional, managerial, skilled non-manual, skilled manual, and semiskilled/unskilled manual. We grouped semi-skilled and unskilled into one category due to small numbers in these categories. If either the mother or her partner had missing social class data or were a single parent household, we used the available parental occupation position.

Highest parental education attainment

We derived highest parental educational attainment from maternal report of her educational attainment and that of her partner at 32 weeks' gestation. Potential responses were based on the Office for National Statistics categorisation: 'O-level/ general certificate of secondary education (GCSE)', 'Advanced-level (A level)', and 'university degree'.  O-level and GCSE indicate secondary school level education and A-levels indicate a subject-based education qualification. O-levels and GCSEs represented compulsory-level schooling from 1976 to 1997 and was coded as 'compulsory education'. If one of the parents' educational attainments is missing or the mother was from a single parent household, we used the available educational attainment.

Financial hardship

At 32 weeks' gestation, mothers were asked "how difficult at the moment do you find it to afford" the following items: food, heating, clothing, rent or mortgage, and things for the baby/child. Possible responses were scored on a four-point Likert scale: "not difficult" (0), "slightly difficult" (1), "fairly difficult" (2), or "very difficult" (3).  We added these individual items' score to derive a continuous total score ranging from 0 to 15 in which higher scores represent greater financial hardship.

Area-level deprivation

Mothers provided residential postcodes at 32 weeks' gestation. These were previously linked to Townsend deprivation index scores,[6] a measure of material deprivation obtained from the 1991 Census data for enumeration district.[7] Townsend index scores are calculated using standardised values of four indicators capturing percentage of: (i) households without a car, (ii) households who do not own their home; (iii) people aged 16 years or over who are economically inactive, and (iv) overcrowded households. We used the continuous z-scores where higher scores indicate higher levels of deprivation.

### Direct Acyclic Graphs

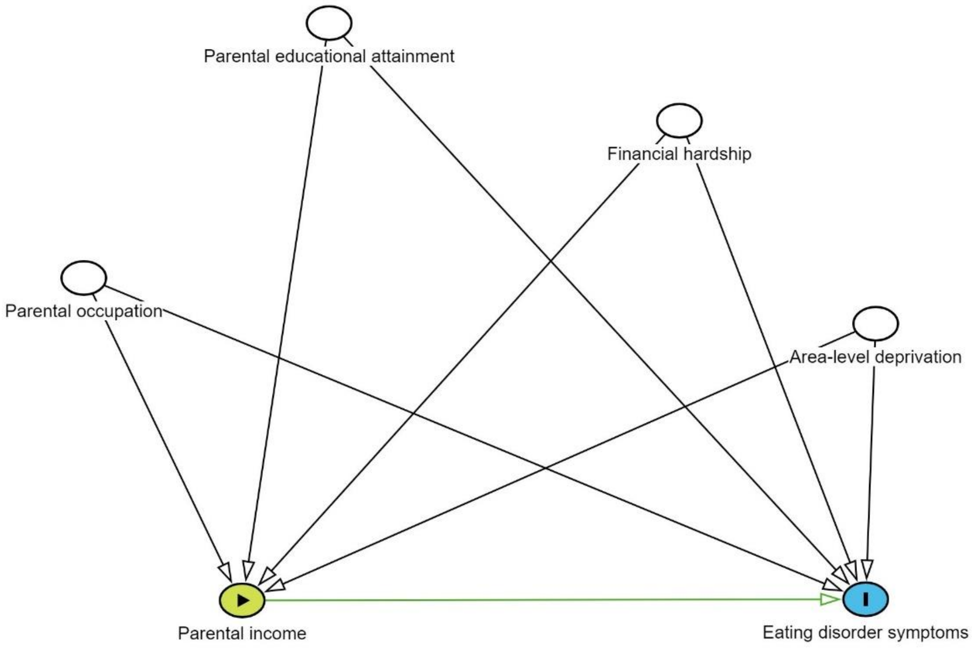

Figure S1: Direct Acyclic Graph hypothesizing relationship between socioeconomic indicators for our main analysis

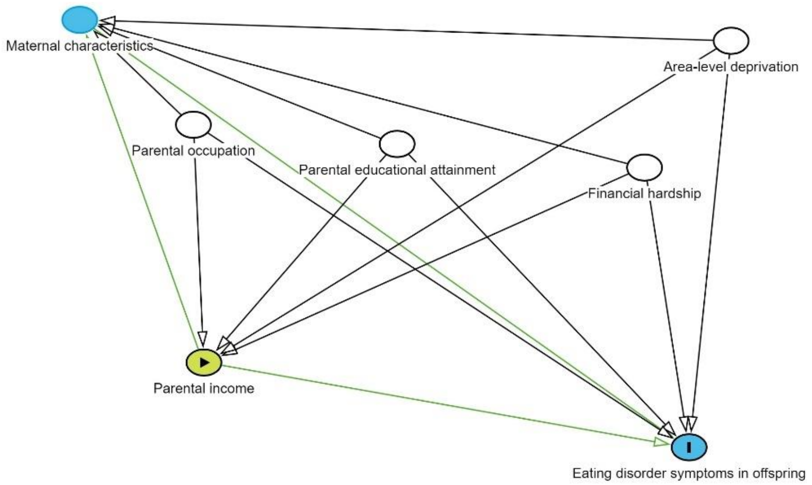

Figure S2: Direct Acyclic Graph hypothesizing maternal characteristics mediating the relationship between socioeconomic position indicators in childhood and eating disorder symptoms in adolescence.

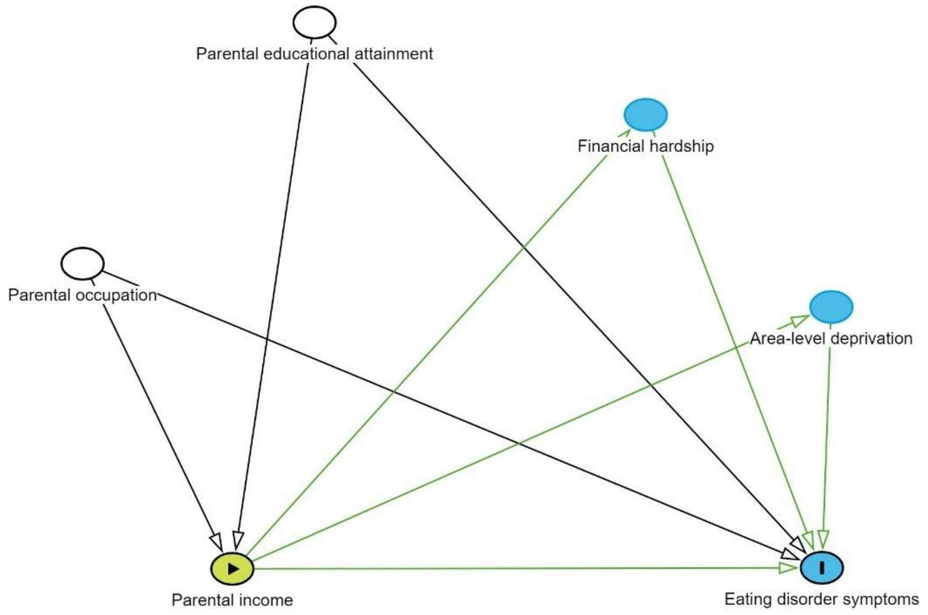

Figure S3: Direct Acyclic Graph hypothesizing different association between socioeconomic position indicators. In this graph, structural indicators (education and occupation) affect maternal indicators (income), which in turn affect perceptual indicators

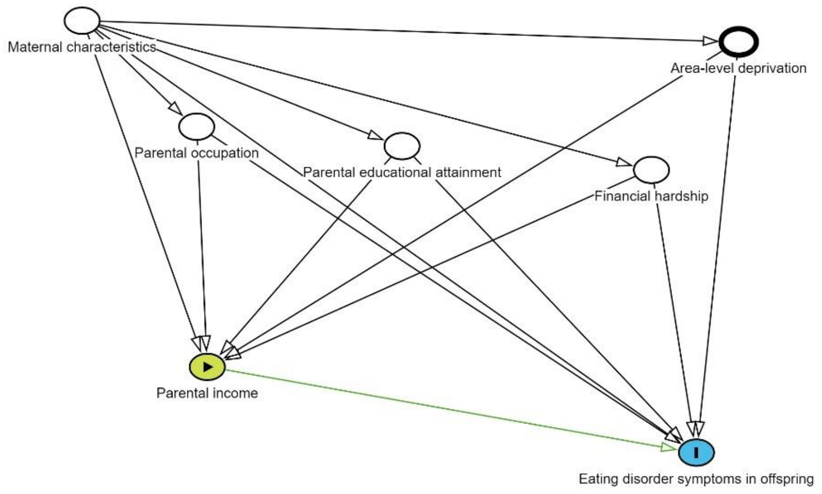

Figure S4: Direct Acyclic Graph hypothesizing maternal characteristics confounding the relationship between socioeconomic position indicators in childhood and eating disorder symptoms in adolescence.

#### Confounders

Information on maternal characteristics were collected through postal questionnaires throughout pregnancy. We used maternal age at birth of study child as a continuous variable. Marital status during pregnancy was asked at 8-week gestations and was coded as a binary variable indicating whether the mother was currently married or unmarried. For maternal depressive symptoms, we used the Edinburgh Post-Natal Depression Scale (EPDS) scale [8] at 12 weeks' of gestation, which ranges from 0 to 30 with higher scores indicating higher levels of depressive symptoms. [8] Maternal lifetime history of eating disorders at 12-week gestation were recoded into a binary variable of indicating whether the mother had previously experienced either an eating disorder (anorexia nervosa, bulimia nervosa, or both) or not. Child ethnicity was coded as a binary measure of “Minortised ethnic group” or “White” and was derived from parent’s response of their ethnicity.

**Deviation from protocol**

We report the following deviations from the protocol published on Open Science Framework (https://osf.io/hsg85/)

| Original protocol | The study |
|---|---|
| The original protocol did not include a sensitivity analysis investigating a potential competing hypothesis that structural socioeconomic indicators (e.g., education and occupation) as well as affecting each other (i.e. education affecting occupation), could affect material indicators (income) which will in turn affect perceptual indicators (financial hardship) | The study included a sensitivity analysis based on the causal assumption that education could affect occupation, education and occupation could affects income, and education, occupation, and income could affect financial hardship. We added this analysis in as we found a competing explanation for our main assumption in the literature. |
| In the protocol, we stated that "Adolescents rated their satisfaction with nine body parts including weight, figure, stomach, waist, thighs, buttocks, hips, and legs. Questions for female adolescents were different from questions for male respondents, with 'body build' in the male adolescent's questionnaire being changed to 'breasts" in the female respondent's questionnaire." | In our study, we stated that we would include "body parts included weight, figure, stomach, waist, thighs, buttocks, hips, legs, breasts/body build (depending on whether the respondent was female or male, respectively), face, and hair." To include all the items of the questionnaire. |
| In the protocol we stated that we would include "other eating style questions" at 21 as an auxiliary variables. | This auxiliary variable was not available in the dataset, therefore it was not included in the imputation model. |
| The original protocol did not include a sensitivity analysis investigating the role of ethnicity as a potential confounder for the association between childhood socioeconomic position and eating disorder symptoms across adolescence. This was because the proportion of ALSPAC participants from ethnic minority backgrounds is very small (<5% in the full sample, and 3.8% among those with complete exposure) | To address any concerns that ethnicity could be a confounder of the association between exposure and outcome in the study we study included a sensitivity analysis to investigate whether adjusting for ethnicity would lead to changes in effect sizes or estimates. |

**Auxiliary variables**

Auxiliary variables included child's depressive symptoms at age 13, 14, and 18 years old measured with the Moods and Feelings Questionnaire, [9] child's internalising and externalising symptoms at ages 7, 9, and 11 years reported by the mother using the Strengths and Difficulties Questionnaire [10], child's total, verbal, and performance IQ at age 8 years, maternal smoking in pregnancy at 18 weeks' of gestation, and maternal pre-pregnancy body dissatisfaction at 18 weeks' of gestation, child's BMI at 7, 9, 10, 11, 14, 16, 18 years old, eating behaviours using the Dutch Eating Behaviour Questionnaire (total of the restrictive, emotional, and external eating scales) [11] at 14 years old, bullying at 8 and 10 years old, autistic traits measured by the Social and Communication Disorders Checklist [12] at 7, 11, 14, 16 years old.

Table S1: Characteristics of respondents with at least one available outcome measurement or no available data on eating disorder outcomes among participants with complete exposure data (N=7,824)

| | Disordered eating behaviours | | Weight and shape concerns | | Body dissatisfaction | |
|---|---|---|---|---|---|---|
| **Parental socioeconomic position** | No available measurements | At least one available measurement at age 14, 16, 18 years | No available measurements | At least one available measurement at age 14, 18 years | No available measurements | Available body dissatisfaction data at age 14 years |
| | **n (%)** | **n (%)** | **n (%)** | **n (%)** | **n (%)** | **n (%)** |
| **Total** | 2,802 (35.8) | 5,022 (64.2) | 3,133 (40.0) | 4,691 (60.0) | 6,838 (87.4) | 986 (12.6) |
| **Highest parental education** | | | | | | |
| University degree | 434 (22.2) | 1520 (77.8) | 522 (26.7) | 1432 (73.3) | 1647 (84.3) | 307 (15.7) |
| A-level | 935 (34.1) | 1808 (65.9) | 1047 (38.2) | 1696 (61.8) | 2392 (87.2) | 351 (12.8) |
| Compulsory education | 1433 (45.8) | 1694 (54.2) | 1564 (50.0) | 1563 (50.0) | 2799 (89.5) | 328 (10.5) |
| **Highest parental occupation** | | | | | | |
| Professional | 260 (22.5) | 893 (77.5) | 305 (26.5) | 848 (73.6) | 967 (83.9) | 186 (16.1) |
| Managerial | 1108 (32.5) | 2296 (67.5) | 1268 (37.3) | 2136 (62.8) | 2964 (87.1) | 440 (12.9) |
| Skilled non-manual | 798 (40.0) | 1198 (60.0) | 874 (43.8) | 1122 (56.2) | 1770 (88.7) | 226 (11.3) |
| Skilled manual | 430 (48.7) | 453 (51.3) | 465 (52.7) | 418 (47.3) | 780 (88.3) | 103 (11.7) |
| Semi-skilled/unskilled manual | 206 (53.1) | 182 (46.9) | 221 (57.0) | 167 (43.0) | 357 (92.0) | 31 (8.0) |
| **Fifths of equivalised parental income** | | | | | | |
| Highest 20% | 451 (26.5) | 1253 (73.5) | 525 (30.8) | 1179 (69.2) | 1446 (84.9) | 257 (15.1) |
| 2 | 513 (30.7) | 1157 (69.3) | 587 (35.2) | 1083 (64.8) | 1429 (85.6) | 241 (14.4) |
| 3 | 579 (37.1) | 981 (62.9) | 650 (41.7) | 910 (58.3) | 1361 (87.2) | 199 (12.8) |
| 4 | 609 (39.7) | 926 (60.3) | 663 (43.2) | 872 (56.8) | 1375 (89.6) | 160 (10.4) |
| Lowest 20% | 650 (48.0) | 705 (52.0) | 708 (52.3) | 647 (47.8) | 1226 (90.5) | 129 (9.5) |
| **Financial hardship*** | | | | | | |
| No (<5) | 1981 (33.1) | 4,000 (66.9) | 2245 (37.5) | 3736 (62.5) | 5189 (86.8) | 792 (13.2) |
| Yes (≥5) | 821 (44.6) | 1,022 (55.4) | 888 (48.2) | 955 (51.8) | 1649 (89.5) | 194 (10.5) |

For descriptive table purposes we defined: [a] Experiencing high financial hardship as scoring 5 or above (75th percentile of scores of the total sample) on the financial hardship scale. [b] High area-level deprivation, as having a standardised Townsend score equal or lower than 0.36 which was the average deprivation score in the UK in 1990. Details on maternal characteristics: [c] Maternal report at 12 weeks of pregnancy.[d] Maternal report at 8 weeks of pregnancy. [e] Total Edinburgh Postnatal Depression Scale score at 12 weeks of pregnancy

Table S1 continued

| | Disordered eating behaviour | Weight and shape concerns | Body dissatisfaction |
|---|---|---|---|

| Parental socioeconomic position | No available measurements | At least one available measurement at age 14, 16, 18 years | No available measurements | At least one available measurement at age 14, 16, and 18 years | No available measurements | Available body dissatisfaction data at 14 years |
|---|---|---|---|---|---|---|
| | **n (%)** | **n (%)** | **n (%)** | **n (%)** | **n (%)** | **n (%)** |
| **Standardised area-level deprivation scores[b]** | | | | | | |
| High area level deprivation | 916 (44.4) | 1146 (55.6) | 995 (48.3) | 1067 (51.8) | 1838 (89.1) | 224 (10.9) |
| Low area level deprivation | 1886 (32.7) | 3876 (67.3) | 2138 (37.1) | 3624 (62.9) | 5000 (86.8) | 762 (13.2) |
| **Maternal history of eating disorders** | | | | | | |
| Has not experienced eating disorders | 2615 (35.3) | 4796 (64.7) | 2927 (39.5) | 4484 (60.5) | 6474 (97.4) | 937 (12.6) |
| Has experienced eating disorders | 101 (36.5) | 176 (63.5) | 117 (42.2) | 160 (57.8) | 240 (86.6) | 37 (13.4) |
| **Marital status** | | | | | | |
| Not married | 672 (43.9) | 858 (56.1) | 731 (47.8) | 799 (52.2) | 1,369 (89.5) | 161 (10.5) |
| Married | 2077 (33.5) | 4,131 (66.5) | 2,343 (37.7) | 3,865 (62.3) | 5,389 (85.8) | 819 (13.2) |
| | **Mean (SD)** | **Mean (SD)** | **Mean (SD)** | **Mean (SD)** | **Mean (SD)** | **Mean (SD)** |
| **Maternal age at birth of study child** | 27.8(4.7) | 29.2 (4.4) | 27.9 (4.7) | 29.2 (4.4) | 28.6 (4.6) | 20.1 (4.4) |
| **Maternal depressive symptoms** | 7.1 (4.8) | 6.3 (4.5) | 7.1 (4.8) | 6.3 (4.5) | 6.7 (4.7) | 6.2 (4.4) |

For descriptive table purposes we defined: [a] Experiencing high financial hardship as scoring 5 or above (75$^{th}$ percentile of scores of the total sample) on the financial hardship scale. [b] High area-level deprivation, as having a standardised Townsend score equal or lower than 0.36 which was the average deprivation score in the UK in 1990. Details on maternal characteristics: [c] Maternal report at 12 weeks of pregnancy.[d] Maternal report at 8 weeks of pregnancy. [e] Total Edinburgh Postnatal Depression Scale score at 12 weeks of pregnancy

Unconditional model

Table S2: Results of model 1 exploring the association between eating disorder symptoms and age (14-18 years old). Sample based on participants with complete parental socioeconomic data and imputed eating disorder symptoms for those with at least one available eating disorder symptom outcome (N=7824)

| | Disordered eating behaviour | Binge eating | Restrictive eating | Purging | Weight and shape concerns |
|---|---|---|---|---|---|
| | OR (95% CI), p value | OR (95% CI), p value | OR (95% CI), p value | OR (95% CI), p value | Mean difference (95% CI), p value |
| Age | 1.37 (1.30 to 1.44), <.0001 | 1.49 (1.40 to 1.59), <.0001 | 1.19 (1.11 to 1.27), <.0001 | 1.53 (1.34 to 1.74), <.0001 | 0.09 (0.07 to 0.11), <.0001 |

Table S3: Results of model 2 exploring the association between eating disorder symptoms, age (14-18 years old), and age$^2$ Sample based on participants with complete parental socioeconomic data and imputed eating disorder symptoms for those with at least one available eating disorder symptom outcome (N=7824)

| | Disordered eating behaviour | Binge eating | Restrictive eating | Purging |
|---|---|---|---|---|
| | OR (95% CI), p value | OR (95% CI), p value | OR (95% CI), p value | OR (95% CI), p value |
| Age | 1.39 (1.31 to 1.47), <.0001 | 1.55 (1.44 to 1.69), <.0001 | 1.20 (1.12 to 1.28), <.0001 | 1.62 (1.39 to 1.90), <.0001 |
| Age$^2$ | 0.92 (0.89 to 0.96), <.0001 | 0.89 (0.83 to 0.95), 0.001 | 0.95 (0.91 to 0.99), 0.028 | 0.86 (0.79 to 0.95), 0.002 |

Supplementary analyses: Interaction between parental socioeconomic position and age and its association with disordered eating behaviour of adolescents at age 14, 16, and 18 years

Table S4: Stratified odds ratio of adolescent disordered eating behaviour at age 14, 16, and 18 according to parental soceioconomic indicators. Sample based on participants with complete parental socioeconomic data and imputed eating disorder outcomes (N = 7,824)

| | | | **Disordered eating behaviour** |
|---|---|---|---|
| Age | 14 | 16 | 18 |
| Parental socioeconomic position indicator | OR (95% CI) | OR (95% CI) | OR (95% CI) |
| **Fifths of equivalised family income** | | | |
| Highest 20% | Ref | Ref | Ref |
| 2 | 0.93 (0.66 to 1.32) | 0.82 (0.64 to 1.04) | 0.93 (0.72 to 1.18) |
| 3 | 1.11 (0.66 to 1.32) | 0.77 (0.58 to 1.02) | 0.79 (0.61 to 1.03) |
| 4 | 1.12 (0.76 to 1.65) | 0.91 (0.69 to 1.19) | 0.82 (0.63 to 1.07) |
| Lowest 20% | 1.03 (0.71 to 1.51) | 0.75 (0.55 to 1.02) | 0.78 (0.57 to 1.07) |

We found moderate evidence ($p = 0.032$) of an interaction between age and parental income. On the other hand, there was no evidence of an interaction between age$^2$ and any socioeconomic indicators. The age-stratified analyses presented in Table S9 suggests that children with parents in lower income categories have higher odds of 1.12 (95% CI 0.76 to 1.65) for developing an eating disorder compared to the highest parental income category at the age of 14. However, this association reverses by age 16. Those in the lower income category has lower odds of 0.82 (95% CI 0.63 to 1.07) of developing an eating disorder compared to parents of the highest income category.

Supplementary analyses: Association between parental socioeconomic position and binge eating, restrictive eating, and purging in adolescents at age 14, 16, and 18 years

Table S5: Multilevel logistic and linear regression models for binge eating, restrictive eating, and purging and their association with parental socioeconomic position at 14, 16, and 18. Sample based on participants with complete parental socioeconomic data and imputed eating disorder symptoms for those imputed eating disorder outcomes (N=7,824)

| | Binge eating | | Restrictive eating | | Purging | |
|---|---|---|---|---|---|---|
| Parental socioeconomic position indicators | Univariable model Odds ratio (95% CI), p | Multivariable model Odds ratio (95% CI), p | Univariable model Odds ratio (95% CI), p | Multivariable model Odds ratio (95% CI), p | Univariable model Odds ratio (95% CI), p | Multivariable model Odds ratio (95% CI), p |
| **Highest parental education** | | | | | | |
| University degree | Ref | Ref | Ref | Ref | Ref | Ref |
| A -level | 1.35 (0.96 to 1.90), 0.088 | 1.41 (0.95 to 2.10), 0.083 | 1.32 (1.03 to 1.69), 0.028 | 1.22 (0.93 to 1.60), 0.147 | 1.07 (0.66 to 1.83), 0.721 | 0.97 (0.54 to 1.74), 0.915 |
| O-level/GCSE | 1.48 (1.07 to 2.05), 0.020 | 1.46 (0.93 to 2.30), 0.103 | 1.84 (1.45 to 2.32), <.001 | 1.63 (1.21 to 1.20), 0.001 | 1.76 (1.06 to 3.00), 0.037 | 1.42 (0.78 to 2.72), 0.278 |
| **Highest parental occupation** | | | | | | |
| Professional | Ref | Ref | Ref | Ref | Ref | Ref |
| Managerial | 1.17 (0.80 to 1.71), 0.421 | 0.95 (0.63 to 1.44), 0.815 | 1.31 (1.00 to 1.70), 0.046 | 1.11 (0.84 to 1.48), 0.460 | 1.52 (0.85 to 2.72), 0.161 | 1.34 (0.73 to 2.46), 0.350 |
| Skilled non-manual | 1.22 (0.81 to 1.84), 0.332 | 0.86 (0.52 to 1.42), 0.547 | 1.67 (1.27 to 2.22), <.001 | 1.27 (0.90 to 1.80), 0.170 | 1.67 (0.83 to 3.33), 0.147 | 1.22 (0.57 to 2.60), 0.603 |
| Skilled manual | 1.18 (0.70 to 2.00), 0.529 | 0.73 (0.39 to 1.36), 0.324 | 1.72 (1.19 to 2.47), 0.004 | 1.25 (0.80 to 1.95), 0.323 | 1.61 (0.73 to 3.57), 0.238 | 1.06 (0.43 to 2.63), 0.904 |
| Semi-skilled/unskilled | 1.48 (0.73 to 2.91), 0.283 | 0.84 (0.39 to 1.80), 0.651 | 2.15 (1.27 to 3.65), 0.005 | 1.49 (0.84 to 2.68), 0.172 | 3.00 (1.13 to 7.97), 0.028 | 1.84 (0.66 to 5.14), 0.245 |

Tables S5 continued

| Parental socioeconomic position indicators | Binge eating: Univariable model OR (95% CI), p | Binge eating: Multivariable model OR (95% CI), p | Restrictive eating: Univariable model OR (95% CI), p | Restrictive eating: Multivariable model OR (95% CI), p | Purging: Univariable model OR (95% CI), p | Purging: Multivariable model OR (95% CI), p |
|---|---|---|---|---|---|---|
| **Equivalised family income** | | | | | | |
| Highest 20% | Ref | Ref | Ref | Ref | Ref | Ref |
| 2 | 0.88 (0.59 to 1.30),0.513 | 0.78 (0.52 to 1.18), 0.238 | 1.09 (0.83 to 1.43), 0.544 | 0.91 (0.70 to 1.20), 0.517 | 0.85 (0.46 to 1.57), 0.598 | 0.71 (0.38 to 1.29), 0.256 |
| 3 | 0.86 (0.55 to 1.33), 0.489 | 0.70 (0.44 to 1.13), 0.141 | 1.18 (0.88 to 1.57), 0.276 | 0.85 (0.62 to 1.16), 0.296 | 1.41 (0.72 to 2.78), 0.315 | 0.98 (0.50 to 1.94), 0.957 |
| 4 | 1.27 (0.84 to 1.93), 0.261 | 0.96 (0.61 to 1.53), 0.871 | 1.28 (0.96 to 1.71), 0.097 | 0.83 (0.60 to 1.14), 0.251 | 1.58 (0.83 to 2.98), 0.160 | 0.91 (0.46 to 1.77), 0.773 |
| Lowest 20% | 1.35 (0.89 to 2.04), 0.163 | 0.91 (054 to 1.54), 0.723 | 1.25 (0.91 to 1.72), 0.169 | 0.71 (0.50 to 1.01), 0.059 | 1.03 (0.49 to 2.16), 0.946 | 0.45 (0.20 to 1.03), 0.060 |
| **Financial hardship score** | 1.06 (1.02 to 1.10), 0.003 | 1.05 (1.00 to 1.10), 0.036 | 1.06 (1.03 to 1.09), <.001 | 1.05 (1.02 to 1.08), 0.003 | 1.08 (1.02 to 1.15) 0.013 | 1.07 (1.00 to 1.14) 0.036 |
| **Standardised Area-level deprivation score*** | 1.06 (1.01 to 1.12), 0.012 | 1.05 (0.99 to 1.11), 0.062 | 1.02 (0.97 to 1.06), 0.216 | 1.00 (0.97 o 1.04), 0.841 | 1.11 (1.04 to 1.20), 0.003 | 1.10 (1.03 to 1.19), 0.009 |

Sensitivity analysis: partially adjusted analyses

Table S6 Multilevel logistic and linear regression models for disordered eating behaviours and weight and shape concerns at age 14, 16, and 18 according to parental socioeconomic position. Sample based on participants with complete parental socioeconomic data and imputed eating disorder outcomes (N = 7824)

| | | | **Disordered eating behaviour** | | | **Weight and shape conce** |
|---|---|---|---|---|---|---|
| **Parental socioeconomic position** | Adjusted for education<br>OR (95% CI), p | Adjusted for education, occupation<br>OR (95% CI), p | Adjusted for education, occupation, and income<br>OR (95% CI), p | Adjusted for education<br>Mean difference (95% CI), p | Adjusted for education, occupation<br>Mean difference (95% CI), p | Adjuste educa occupation, inc<br>Mean differe (95% C |
| **Highest parental occupation** | | | | | | |
| Professional | Ref | | | Ref | | |
| Managerial | 1.05 (0.81 to 1.36), 0.699 | | | 0.09 (-0.02 to 1.20), 0.102 | | |
| Skilled non-manual | 1.12 (0.81 to 1.55), 0.490 | | | 0.10 (-0.02 to 0.22), 0.095 | | |
| Skilled manual | 1.09 (0.72 to 1.64), 0.678 | | | 0.09 (-0.09 to 0.26), 0.333 | | |
| Semi-skilled/unskilled | 1.43 (0.86 to 2.39), 0.167 | | | 0.15 (-0.10 to 0.39), 0.245 | | |

Table S6 continued

| **Parental socioeconomic position** | | | **Disordered eating behaviour** | | | **Weight and shape conce[illegible]** |
|---|---|---|---|---|---|---|
| | Adjusted for education<br>OR (95% CI), p | Adjusted for education, occupation<br>OR (95% CI), p | Adjusted for education, occupation, and income<br>OR (95% CI), p | Adjusted for education<br>Mean difference (95% CI), p | Adjusted for education, occupation<br>Mean difference (95% CI), p | Adjuste[illegible] educa[illegible] occupation, [illegible] inc[illegible]<br>Mean differe[illegible] (95% C[illegible] |
| **Equivalised income** | | | | | | |
| Highest 20% | Ref | Ref | | Ref | Ref | |
| 2 | 0.89 (0.68 to 1.16), 0.393 | 0.89 (0.68 to 1.16), 0.368 | | 0.01 (-0.90 to 0.11), 0.836 | 0.003 (-0.10 to 0.10), 0.940 | |
| 3 | 0.87 (0.66 to 1.16), 0.343 | 0.86 (0.65 to 1.14), 0.293 | | 0.05 (-0.07 to 0.17), 0.440 | 0.04 (-0.09 to 0.16), 0.552 | |
| 4 | 1.05 (0.80 to 1.38), 0.705 | 1.03 (0.78 to 1.35), 0.859 | | 0.007 (-0.05 to 0.19), 0.266 | 0.06 (-0.07 to 0.18), 0.374 | |
| Lowest 20% | 1.01 (0.74 to 1.38), 0.954 | 0.97 (0.71 to 1.33), 0.845 | | 0.02 (-0.11 to 0.16), 0.723 | 0.01 (-0.13 to 0.14), 0.903 | |
| | Coefficient (95% CI), p | Coefficient (95% CI), p | Coefficient (95% CI), p | Coefficient (95% CI), p | Coefficient (95% CI), p | Coeffic[illegible] (95% C[illegible] |
| **Financial hardship scores** | 1.06 (1.03 to 1.08), <.001 | 1.06 (1.03 to 1.09), <.001 | 1.06 (1.03 to 1.09), <.001 | 0.02 (0.01 to 0.04), 0.002 | 0.02 (0.01 to 0.04), 0.003 | 0.02 (0.01 to 0[illegible] 0[illegible] |

Table S7: Linear regression model for body dissatisfaction and weight and shape concerns at age 14, 16, and 18 according to parental socioeconomic position. Sample based on participants with complete parental socioeconomic data and imputed eating disorder outcomes (N = 7824)

| | | | **Body dissatisfaction** |
|---|---|---|---|
| **Parental socioeconomic position indicators** | Adjusted for education Mean differences (95% CI), p-value | Adjusted for education, occupation Mean differences (95% CI), p-value | Adjusted for education, occupation, and income |
| **Highest parental occupation** | | | |
| Professional | Ref | Ref | |
| Managerial | 0.14 (-1.36 to 1.63), 0.857 | | |
| Skilled non-manual | 0.34 (-1.35 to 2.02), 0.692 | | |
| Skilled manual | 1.00 (-1.43 to 3.42), 0.415 | | |
| Semi-skilled/unskilled | 0.34 (-2.66 to 3.35), 0.820 | | |
| **Fifths of equivalised family income** | | | |
| Highest 20% | Ref | Ref | |
| 2 | 0.49 (-0.97 to 1.94), 0.506 | 0.48 (-0.99 to1.94), 0.520 | |
| 3 | -0.61 (-2.43 to 1.22), 0.510 | -0.66 (-2.51 to 1.20), 0.483 | |
| 4 | 1.86 (0.03 to 3.68), 0.047 | 1.77 (-0.08 to 3.62), 0.061 | |
| Lowest 20% | 0.30 (-1.66 to 2.26), 0.760 | 0.20 (-1.80 to 2.20), 0.845 | |
| | Coefficient (95% CI), p | Coefficient (95% CI), p | Coefficient (95% CI), p |
| **Financial hardship score** | 0.22 (0.07 to 0.38), 0.005 | 0.22 (0.06 to 0.38), 0.007 | 0.22 (0.06 to 0.38), 0.009 |

Sensitivity analysis: adjusting for maternal characteristics

Table S8: Multilevel logistic and linear regression models adjusted for maternal characteristics for disordered eating behaviours at age 14, 16, and 18 according to parental socioeconomic position. Sample based on participants with complete parental socioeconomic data, imputed eating disorder symptoms for those with at least one available eating disorder symptom outcome, and imputed confounders (N = 7824)

| **Parental socioeconomic position** | **Any behavioural eating disorder symptoms** | | **Weight and shape concer[illegible]** | |
|---|---|---|---|---|
| | 1: Adjusted for maternal characteristics<br>Odds ratios (95% CI), p-value | 2: 1+ adjusted for socioeconomic position indicator<br>Odds ratios (95% CI), p-value | 1: Adjusted for maternal characteristics<br>Mean difference (95% CI), p | 2: 1+ adjusted f[illegible] socioeconomic positi[illegible] indicat[illegible]<br>Mean differen[illegible] (95% CI),[illegible] |
| **Highest parental education** | | | | |
| University degree | Ref | Ref | Ref | R[illegible] |
| A -level | 1,25 (1.01 to 1.56), 0.041 | 1.27 (0.99 to 1.61) , 0.051 | 0.06 (-0.03 to 0.15), 0.180 | 0.03 (-0.07 to 0.12), 0.5[illegible] |
| O-level/GCSE | 1.57 (1.25 to 1.97), <.001 | 1.56 (1.17 to 2.07), 0.002 | 0.07 (-0.03 to 0.17), 0.154 | 0.03 (-0.09 to 0.15), 0.6[illegible] |
| **Highest parental occupation** | | | | |
| Professional | Ref | Ref | Ref | R[illegible] |
| Managerial | 1.18 (0.93 to 1.49), 0.181 | 1.03 (0.79 to 1.33), 0.846 | 0.10 (-0.001 to 0.19), 0.054 | 0.07 (-0.03 to 0.19), 0.1[illegible] |
| Skilled non-manual | 1.33 (1.00 to 1.77), 0.050 | 1.05 (0.76 to 1.47), 0.760 | 0.11 (0.01 to 0.21), 0.037 | 0.08 (-0.04 to 0.20), 0.2[illegible] |
| Skilled manual | 1.25 (0.87 to 1.79), 0.219 | 0.94 (0.62 to 1.43), 0.772 | 0.09 (-0.07 to 0.25), 0.289 | 0.04 (-0.14 to 0.22), 0.6[illegible] |
| Semi-skilled/unskilled | 1.63 (1.01 to 2.64), 0.047 | 1.18 (0.70 to 1.98), 0.532 | 0.14 (-0.09 to 0.37), 0.241 | 0.09 (-0.16 to 0.34), 0.4[illegible] |
| **Fifths of equivalised family income** | | | | |
| Highest 20% | Ref | Ref | Ref | R[illegible] |
| 2 | 0.94 (0.73 to 1.23), 0.663 | 0.84 (0.64 to 1.09), 0.191 | 0.01 (-0.09 to 0.11), 0.775 | -0.02 (-0.12 to 0.09), 0.7[illegible] |
| 3 | 0.97 (0.74 to 1.28), 0.840 | 0.78 (0.59 to 1.03), 0.082 | 0.06 (-0.05 to 0.17), 0.323 | 0.01 (-0.12 to 0.13), 0.9[illegible] |
| 4 | 1.17 (0.90 to 1.51), 0.230 | 0.87 (0.66 to 1.14), 0.306 | 0.07 (-0.04 to 0.18), 0.213 | -0.002 (-0.13 to 0.12), 0.9[illegible] |
| Lowest 20% | 1.06 (0.78 to 1.42), 0.720 | 0.71 (0.52 to 0.98), 0.039 | 0.01 (-0.11 to 0.13), 0.898 | -0.09 (-0.23 to 0.05), 0.2[illegible] |
| | OR (95% CI), p | OR (95% CI), p | Coefficient (95% CI), p | Coefficient (95% CI),[illegible] |
| **Financial hardship score** | 1.05 (1.02 to 1.08), <.001 | 1.05 (1.02 to 1.08), 0.001 | 0.02 (0.003 to 0.03), 0.019 | 0.02 (0.002 to 0.03), 0.0[illegible] |
| **Standardised Area-level deprivation score** | 1.03 (0.99 to 1.07), 0.097 | 1.02 (0.99 to 1.06), 0.190 | 0.01 (-0.003 to 0.02), 0.117 | 0.01 (-0.004 to 0.02), 0.1[illegible] |

Table S9: Linear regression models adjusted for maternal characteristics for body dissatisfaction at age 14 according to socioeconomic indicators. Sample based on participants with complete parental socioeconomic data, imputed eating disorder symptoms for those with at least one available eating disorder symptom outcome, and imputed confounders (N = 7,824)

| | Body dissatisfaction | |
|---|---|---|
| | 1: Adjusted for maternal characteristics | 2: 1+ adjusted for socioeconomic position indicator |
| Parental socioeconomic position indicators | Mean differences (95% CI), p-value | Mean differences (95% CI), p-value |
| **Highest parental education** | | |
| University degree | Ref | Ref |
| A -level | 0.12 (-1.07 to 1.31), 0.841 | -0.005 (--1.22 to 1.21), 0.994 |
| O-level/GCSE | 0.82 (-0.36 to 2.00), 0.171 | 0.58 (-0.76 to 1.92), 0.394 |
| **Highest parental occupation** | | |
| Professional | Ref | Ref |
| Managerial | 0.17 (-1.22 to 1.57), 0.804 | -0.03 (-1.57 to 1.51), 0.966 |
| Skilled non-manual | 0.50 (-1.06 to 2.06), 0.526 | 0.72 (-1.71 to 1.85), 0.936 |
| Skilled manual | 1.06 (-1.21 to 3.34), 0.354 | 0.40 (-2.11 to 2.92), 0.750 |
| Semi-skilled/unskilled | 0.37 (-2.55 to 2.39), 0.801 | -0.39 (-3.54 to 2.77), 0.806 |
| **Equivalised family income** | | |
| Highest 20% | Ref | Ref |
| 2 | 0.48 (-0.92 to 1.88), 0.495 | 0.03 (-1.17 to 1.74), 0.699 |
| 3 | -0.56 (-2.30 to 1.17), 0.521 | -0.96 (-2.80 to 0.88), 0.300 |
| 4 | 1.84 (0.10 to 3.60), 0.039 | 1.22 (-0.62 to 3.07), 0.190 |
| Lowest 20% | 0.13 (-1.73 to 1.99), 0.887 | -0.73 (-2.80 to 1.35), 0.488 |
| | Coefficient (95% CI), p | Coefficient (95% CI), p |
| **Financial hardship score** | 0.17 (0.003 to 0.33), 0.045 | 0.15 (-0.02 to 0.32), 0.087 |
| **Standardised Area-level deprivation score** | 0.09 (-0.09 to 0.26), 0.340 | 0.06 (-0.12 to 0.24), 0.522 |

Sensitivity analysis: completed case analysis

Table S10: Multilevel logistic and linear regression models for disordered eating behaviours and its association to parental socioeconomic position at age 14, 16, and 18. Sample based on participants with complete parental socioeconomic data and at least one available eating disorder symptom outcome in non-imputed data

| | **Disordered eating behaviour (n=5,022)** | | **Weight and shape concerns (n=4,69[illegible]** | |
|---|---|---|---|---|
| **Parental socioeconomic position** | Univariable model Odds ratios (95% CI), p-value | Multivariable model Odds ratios (95% CI), p-value | Univariable model Mean difference (95% CI), p | Multivariable mo[illegible] Mean differen[illegible] (95% CI),[illegible] |
| **Highest parental education** | | | | |
| University degree | Ref | Ref | Ref | R[illegible] |
| A -level | 1.24 (0.97 to 1.57) | 1.21 (0.92 to 1.59) | 0.05 (-0.03 to 0.13) | 0.01 (-0.08 to 0.1[illegible] |
| O-level/GCSE | 1.63 (1.28 to 2.07) | 1.48 (1.10 to 2.00) | 0.05 (-0.03 to 0.13) | -0.003 (-0.11 to 0.1[illegible] |
| | p=0.003 | p=0.036 | P=0.435 | p=0.9[illegible] |
| **Highest parental occupation** | | | | |
| Professional | Ref | Ref | Ref | R[illegible] |
| Managerial | 1.12 (0.86 to 146) | 0.99 (0.74 to 1.33) | 0.10 (0.01 to 0.19) | 0.10 (-0.0002 to 0.2[illegible] |
| Skilled non-manual | 1.46 (1.08 to 1.96) | 1.14 (0.80 to 1.63) | 0.11 (0.01 to 0.21) | 0.11 (-0.02 to 0.2[illegible] |
| Skilled manual | 1.53 (1.04 to 2.26) | 1.08 (0.69 to 1.69) | 0.11 (-0.03 to 0.24) | 0.09 (-0.07 to 0.2[illegible] |
| Semi-skilled/unskilled | 1.51 (0.86 to 2.63) | 1.00 (0.56 to 1.85) | 0.05 (-0.15 to 0.25) | 0.03 (-0.18 to 0.2[illegible] |
| | p=0.036 | p=0.869 | p=0.186 | p=0.3[illegible] |
| **Equivalised family income** | | | | |
| Highest 20% | Ref | Ref | Ref | R[illegible] |
| 2 | 0.96 (0.73 to 1.27) | 0.83 (0.63 to 1.10) | 0.02 (-0.07 to 0.12) | -0.01 (-0.11 to 0.0[illegible] |
| 3 | 1.04 (0.78 to 1.39) | 0.78 (0.57 to 1.06) | 0.06 (-0.04 to 0.16) | -0.01 (-0.11 to 0.1[illegible] |
| 4 | 1.22 (0.91 to 1.64) | 0.82 (0.59 to 1.13) | 0.03 (-0.07 to 0.13) | -0.06 (-0.17 to 0.0[illegible] |
| Lowest 20% | 1.28 (0.93 to 1.77) | 0.74 (0.51 to 1.08) | -0.001 (-0.11 to 0.11) | -0.13 (-0.26 to 0.00[illegible] |
| | p=0.299 | p=0.483 | p=0.792 | p=0.3[illegible] |
| | Coefficient (95% CI), p | Coefficient (95% CI), p | Coefficient (95% CI), p | Coefficie[illegible] (95% CI),[illegible] |
| **Financial hardship score** | 1.07 (1.04 to 1.10) p<.001 | 1.06 (1.03 to 1.10) p<.001 | 0.02 (0.01 to 0.03) p<.001 | 0.02 (0.01 to 0.0[illegible] p<.0[illegible] |
| **Standardised Area-level deprivation score** | 1.05 (1.02 to 1.09) p=0.004 | 1.04 (0.99 to 1.08) p=0.061 | 0.01 (-0.002 to 0.02) p=0.103 | 0.01 (-0.003 to 0.0[illegible] p=0.1[illegible] |

Table S11: Linear regression models for body dissatisfaction at age 14 according to socioeconomic indicators Sample based on participants with complete parental socioeconomic data and those with completed body dissatisfaction outcomes in non-imputed data (N=986)

| | **Body dissatisfaction** | |
|---|---|---|
| Parental socioeconomic position indicators | Univariable model<br>Mean differences (95% CI), p-value | Multivariable model<br>Mean differences (95% CI), p-value |
| **Highest parental education** | | |
| University degree | Ref | Ref |
| A -level | 0.29 (-1.29 to 1.87) | 0.20 (-1.62 to 2.01) |
| O-level/GCSE | 1.49 (-0.11 to 3.10) | 1.00 (-1.06 to 3.07) |
| | p=0.147 | p=0.554 |
| **Highest parental occupation** | | |
| Professional | Ref | Ref |
| Managerial | 0.10 (-1.65 to 1.87) | -0.09 (-2.03 to 1.85) |
| Skilled non-manual | 0.70 (-1.29 to 2.69) | 0.18 (-2.23 to 2.58) |
| Skilled manual | 3.59 (1.12 to 6.06) | 2.65 (-0.31 to 5.60) |
| Semi-skilled/unskilled | -0.55 (-4.46 to 3.35) | -1.93 (-6.21 to 2.35) |
| | p=0.030 | p=0.130 |
| **Fifths of equivalised family income** | | |
| Highest 20% | Ref | Ref |
| 2 | -0.27 (-2.08 to 1.54) | -0.78 (-2.64 to 1.08) |
| 3 | -0.79 (-2.69 to 1.11) | -1.96 (-4.02 to 0.09) |
| 4 | 2.05 (0.02 to 4.08) | 0.21 (-2.10 to 2.51) |
| Lowest 20% | 0.33 (-1.85 to 2.50) | -1.71 (-4.22 to 0.80) |
| | p=0.106 | p=0.179 |
| | Coefficient (95% CI), p value | Coefficient (95% CI), p value |
| **Financial hardship score** | 0.33 (0.14 to 0.54) | 0.31 (0.09 to 0.53) |
| | p=0.001 | p=0.005 |
| **Standardised Area-level deprivation score** | 0.13 (-0.10 to 0.37) | 0.04 (-0.21 to 0.29) |
| | p=0.267 | p=0.736 |

**Sensitivity analysis: adjusted for ethnicity**

Table S12: Multilevel logistic and linear regression models for disordered eating behaviours and its association to parental socioeconomic position at age 14, 16, and 18. Sample based on participants with imputed confounders and outcomes and non-imputed ethnicity data (N=7,174)

| | Any behavioural eating disorder symptoms | | | Weight and shape concerns | | |
|---|---|---|---|---|---|---|
| **Parental socioeconomic position** | 1: Univariable model<br>Odds ratio (95% CI), p-value | 2: 1+Adjusted for ethnicity<br>Odds ratios (95% CI), p-value | 3: 2+ adjusted for socioeconomic position indicator<br>Odds ratios (95% CI), p-value | 1: Univariable model<br>Mean difference (95% CI), p-value | 2: 1+Adjusted for ethnicity<br>Mean difference (95% CI), p | 3: 2+ adjusted for socioeconomic position indicator<br>Mean difference (95% CI), p |
| **Highest parental education** | | | | | | |
| University degree | Ref | Ref | Ref | Ref | Ref | Ref |
| A -level | 1.35 (1.09 to 1.69), 0.006 | 1.36 (1.09 to 1.69), 0.006 | 1.31 (1.03 to 1.66), 0.030 | 0.07 (-0.02 to 0.15), 0.111 | 0.07 (-0.01 to 0.15), 0.105 | 0.02 (-0.07 to 0.12), 0.634 |
| O-level/GCSE | 1.80 (1.45 to 2.22), <.0001 | 1.80 (1.46 to 2.23), <.0001 | 1.63 (1.23 to 2.16), 0.001 | 0.09 (-0.002 to 0.18), 0.056 | 0.09 (-0.001 to 0.18), 0.053 | 0.02 (-0.09 to 0.14), 0.697 |
| **Highest parental occupation** | | | | | | |
| Professional | Ref | Ref | Ref | Ref | Ref | Ref |
| Managerial | 1.28 (1.00 to 1.62), 0.047 | 1.28 (1.01 to 1.63), 0.043 | 1.05 (0.81 to 1.36), 0.702 | 0.11 (0.01 to 0.21), 0.025 | 0.11 (0.02 to 0.21), 0.023 | 0.08 (-0.02 to 0.19), 0.129 |
| Skilled non-manual | 1.55 (1.18 to 2.03), 0.002 | 1.56 (1.19 to 2.04), 0.002 | 1.10 (0.80 to 1.53), 0.557 | 0.13 (0.03 to 0.23), 0.010 | 0.13 (0.03 to 0.23), 0.009 | 0.08 (-0.04 to 0.20), 0.178 |
| Skilled manual | 1.07 (1.04 to 1.10), <.0001 | 1.56 (1.09 to 2.22), 0.014 | 1.01 (0.66 to 1.54), 0.976 | 0.12 (-0.04 to 0.27), 0.136 | 0.12 (-0.04 to 0.27), 0.134 | 0.05 (-0.13 to 0.23), 0.597 |
| Semi-skilled/unskilled | 2.10 (1.32 to 3.34), 0.002 | 2.11 (1.33 to 3.34), 0.002 | 1.28 (0.77 to 2.12), 0.333 | 0.17 (-0.06 to 0.40), 0.140 | 0.17 (-0.06 to 0.40), 0.137 | 0.10 (-0.16 to 0.35), 0.450 |

Table S12 continued

| **Parental socioeconomic position** | **Any behavioural eating disorder symptoms** | | | **Weight and shape concerns** | | |
|---|---|---|---|---|---|---|
| | 1: Univariable model<br>Odds ratio (95% CI), p-value | 2: 1+Adjusted for ethnicity<br>Odds ratios (95% CI), p-value | 3: 2+ adjusted for socioeconomic position indicator<br>Odds ratios (95% CI), p-value | 1: Univariable model<br>Mean difference (95% CI), p-value | 2: 1+Adjusted for ethnicity<br>Mean difference (95% CI), p | 3: 2+ adjusted for socioeconomic position indicator<br>Mean difference (95% CI), p |
| **Fifths of equivalised family income** | | | | | | |
| Highest 20% | Ref | Ref | Ref | Ref | Ref | Ref |
| 2 | 1.00 (0.77 to 1.29), 0.980 | 1.00 (0.77 to 1.29), 0.985 | 0.84 (0.65 to 1.10), 0.208 | 0.02 (-0.08 to 0.12), 0.657 | 0.02 (-0.08 to 0.12), 0.650 | -0.02 (-0.12 to 0.08), 0.771 |
| 3 | 1.08 (0.82 to 1.42), 0.595 | 1.08 (0.82 to 1.42), 0.588 | 0.79 (0.59 to 1.05), 0.103 | 0.07 (-0.04 to 0.19), 0.228 | 0.07 (-0.04 to0.19), 0.224 | 0.01 (-0.12 to 0.13), 0.934 |
| 4 | 1.34 (1.03 to 1.74), 0.029 | 1.34 (1.03 to 1.73), 0.029 | 0.87 (0.66 to 1.16), 0.351 | 0.09 (-0.02 to 0.20), 0.091 | 0.09 (-0.02 to 0.20), 0.091 | 0.0001 (-0.12 to 0.12), 0.999 |
| Lowest 20% | 1.34 (1.01 to 1.80), 0.045 | 1.34 (1.00 to 1.79), 0.048 | 0.75 (0.55 to 1.04), 0.086 | 0.06 (-0.07 to 0.18), 0.359 | 0.06 (-0.07 to 0.18), 0.374 | -0.08 (-0.22 to 0.07), 0.302 |
| | Coefficient (95% CI), p | Coefficient (95% CI), p | Coefficient (95% CI), p | Coefficient (95% CI), p | Coefficient (95% CI), p | Coefficient (95% CI), p |
| **Financial hardship score** | 1.07 (1.04 to 1.10), <.0001 | 1.07 (1.04 to 1.10), <.0001 | 1.06 (1.03 to 1.09), <.001 | 0.02 (0.01 to 0.04), 0.001 | 0.02 (0.01 to 0.04), 0.001 | 0.02 (0.01 to 0.04), 0.004 |
| **Standardised Area-level deprivation score** | 1.05 (1.02 to 1.09), 0.002 | 1.05 (1.02 to 1.09), 0.003 | 1.03 (0.99 to 1.07), 0.086 | 0.01 (0.001 to 0.03), 0.039 | 0.01 (0.0002 to 0.03), 0.046 | 0.01 (-0.003 to 0.02), 0.147 |

Table S13: Linear regression models for body dissatisfaction at age 14 according to socioeconomic indicators Sample based on participants with complete parental socioeconomic data and those with completed body dissatisfaction outcomes in non-imputed data (n=7,174)

| | | | **Body dissatisfaction** |
|---|---|---|---|
| | 1:Univariable model | 2: 1+adjusted for ethnicity | 3: 2+adjusted for ethnicity and socioeconomic position |
| Parental socioeconomic position indicators | Mean differences (95% CI), p value | Mean differences (95% CI) p value | Mean differences (95% CI) p value |
| **Highest parental education** | | | |
| University degree | Ref | Ref | Ref |
| A -level | 0.25 (-0.93 to 1.42), 0.679 | 0.26 (-0.92 to 1.43), 0.665 | -0.01 (-1.23 to 1.21), 0.984 |
| O-level/GCSE | 1.07 (-0.09 to 2.24), 0.071 | 1.08 (-0.09 to 2.25), 0.069 | 0.58 (-0.78 to 1.93), 0.401 |
| **Highest parental occupation** | | | |
| Professional | Ref | Ref | Ref |
| Managerial | 0.35 (-1.01 to 1.71), 0.610 | 0.36 (-0.99 to 1.72), 0.596 | 0.04 (-1.50 to 1.57), 0.962 |
| Skilled non-manual | 0.78 (-0.68 to 2.24), 0.292 | 0.79 (-0.67 to 2.26), 0.284 | 0.15 (-1.59 to 1.88), 0.867 |
| Skilled manual | 1.50 (-0.68 to 3.68), 0.175 | 1.50 (-0.67 to 3.68), 0.173 | 0.51 (-2.00 to 3.03), 0.685 |
| Semi-skilled/unskilled | 0.90 (-1.99 to 3.79), 0.537 | 0.91 (-1.98 to 3.80), 0.533 | -0.25 (-3.39 to 2.89), 0.876 |
| **Fifths of equivalised family income** | | | |
| Highest 20% | Ref | Ref | Ref |
| 2 | 0.60 (-0.80 to 2.00), 0.395 | 0.60 (-0.79 to 2.00), 0.392 | 0.30 (-1.16 to 1.76), 0.682 |
| 3 | -0.36 (-2.10 to 1.39), 0.683 | -0.35 (-2.10 to 1.39), 0.688 | -0.95 (-2.80 to 0.90), 0.311 |
| 4 | 2.15 (0.42 to 3.89), 0.016 | 2.15 (0.42 to 3.88), 0.016 | 1.25 (-0.60 to 3.10), 0.682 |
| Lowest 20% | 0.71 (-1.09 to 2.50), 0.435 | 0.69 (-1.11 to 2.49), 0.445 | -0.61 (-2.71 to 1.49), 0.565 |
| | Coefficient (95% CI), p value | Coefficient (95% CI), p value | Coefficient (95% CI), p value |
| **Financial hardship score** | 0.24 (0.09 to 0.39), 0.002 | 0.24 (0.09 to 0.39), 0.002 | 0.21 (0.05 to 0.38), 0.013 |
| **Standardised Area-level deprivation score** | 0.13 (-0.03 to 0.29), 0.115 | 0.13 (-0.04 to 0.29), 0.130 | 0.06 (-0.11 to 0.230, 0.459 |
